# Too much is too much: influence of former stress levels on food craving and weight gain during the COVID-19 period

**DOI:** 10.1101/2022.11.06.22282004

**Authors:** Rachel Granger, Hans P. Kubis

## Abstract

The COVID-19 pandemic and associated social restrictions had an extensive effect on peoples’ lives. Increased rates of weight gain were widely reported, as were declines in the general populations’ mental health, including increases in perceived stress. This study investigated whether higher perceived levels of stress during the pandemic were associated with greater levels of weight gain, and whether poor prior levels of mental health were a factor in higher levels of both stress and weight gain during the pandemic. Underlying changes in eating behaviours and dietary consumption were also investigated. During January-February 2021, UK adults (n=179) completed a self-report online questionnaire to measure perceived levels of stress and changes (current versus pre-COVID-19 restrictions) in weight, eating behaviours, dietary consumption, and physical activity. Participants also reported on how COVID-19 had impacted their lives and their level of mental health prior to the pandemic. Participants with higher levels of stress were significantly more likely to report weight gain and twice as likely to report increased food cravings and comfort food consumption (OR=2.3 and 1.9-2.5, respectively). Participants reporting an increase in food cravings were 6-11 times more likely to snack and to have increased consumption of high sugar or processed foods (OR=6.3, 11.2 and 6.3, respectively). Females reported a far greater number of COVID-19 enforced lifestyle changes and both female gender and having poor mental health prior to the pandemic were significant predictors of higher stress and weight gain during the pandemic. Although COVID-19 and the pandemic restrictions were unprecedented, this study suggests that understanding and addressing the disparity of higher perceived stress in females and individuals’ previous levels of mental health, as well as the key role of food cravings, is key for successfully addressing the continuing societal issue of weight gain and obesity.

## Introduction

After the declaration of the SARS-CoV-2 coronavirus (COVID-19) pandemic in March 2020 [1], restrictions were put in place in many countries around the world, to help curb the spread of the virus [2]. This included the government-imposed restrictions that came into place in the UK on 23rd March 2020 [3]. These initial COVID-19 restrictions caused sudden and radical changes in habits and lifestyles, including self-isolation for individuals more vulnerable to the virus, physical distancing, transfer to home-based working and education, closure of indoor community spaces and sports facilities, use of personal protective equipment (PPE) in work, and job furlough in many sectors [4]. The subsequent lockdowns in Autumn 2020 and Winter 2020-21 (timings varied between countries in the UK) had a significant impact on the UK population [4].

Although COVID-19 restrictions were implemented to improve the health outcome of the pandemic, the high level of hospitalisations and deaths in the UK [5] caused concern both over personal health and the health of loved ones [6]. These concerns, combined with financial insecurity, due to increased job uncertainty and losses [7], and reduced social interactions were linked to a considerable increase on the levels of psychological stress reported by general populations in the UK [6,8] and around the world [9-12].

Since the start of the pandemic there was also an overall reported increase in food intake and weight gain both in the UK and other countries [13-18]. Studies in the first few months of the pandemic reported weight gains in 22-50% of general adult populations [15-18], with follow up studies reporting a substantial subset of people who maintained or reported further weight gain [17,19]. These increases in weight gain were associated with specific dietary behaviours, including increased snacking [14] and greater consumption of processed foods including high sugar and high fat/energy foods [20-22]. This increased weight gain was also often associated with increased levels of food cravings or reductions in food craving control [17,19,20,22,23].

Prior to COVID-19, obesity was already considered to be a global epidemic [24-26]. In addition, people with obesity are likely to suffer more serious health consequences from COVID-19 infection [27-28]. Obesity is a complex multifactorial condition, who’s development is strongly influenced by genetic and behavioural factors, but also by wider socioeconomic and environmental influences of health, which are often linked to higher levels of stress [29]. The level of stress experienced by an individual will often be greater when there is a high perceived level of uncertainty, unpredictability, lack of information and lack of control [30]; all factors that were present during the COVID-19 pandemic.

Prior to the pandemic, long-term chronic stress already had a significant positive association with weight gain [31-34]. Several studies during the pandemic have already reported a link between increased stress and: weight gain [17,19,20,22], changes in eating behaviours [35] or changes in dietary consumption [36].

Food cravings are defined an intense desire to consume a particular food or food type, often in the absence of hunger [37]. Higher levels of food cravings are associated with individuals who have higher BMI/obesity [38] or disordered eating patterns, such as bulimia [20,39]. Higher levels of food cravings or loss of craving control have also been linked to reduced weight loss and early drop-out rates in dietary interventions [39]. Food cravings are generally associated with foods that are high in sugar, fat/energy and are often highly processed [20,23,40]. These foods are also often referred to as comfort foods, as their consumption in excessive amounts is associated with an attempt to sooth emotional upset due to stress, anxiety, depression, loneliness, or anger [41,42].

Often, with current dietary approaches, increased food cravings and excessive comfort eating, which both increase energy intake, have been viewed as drivers of weight gain. [43]. However, some of the main barriers to weight loss and weight maintenance that have been identified in recent studies are mental distress and binge eating (or comfort eating) [44], negative thoughts or moods [45] and traumatic life events [46]. Recent reviews on the role of stress in weight gain has suggested that rather than being viewed as primarily drivers of weight gain, cravings and comfort eating should be considered as mediators of the effect of stress on dietary intake and weight gain [24,32].

The aim of this study was to investigate whether higher levels of stress reported by the general UK population during the pandemic were linked to higher levels of weight gain, and whether these changes were directly associated with increased food cravings and comfort eating. Secondly, we aimed to investigate whether individuals’ level of mental health prior to the pandemic were linked to higher levels of stress and weight gain during the pandemic. In addition, we addressed whether specific lifestyle changes caused by COVID-19 and the UK imposed restrictions were linked to higher levels of stress. We hypothesized that individuals with higher perceived stress would report greater weight gain during the pandemic, with people who had poor mental health before COVID showing the strongest effects. Furthermore, we hypothesized that the weight gain and perceived stress were strongly linked with increased food cravings and comfort food consumption.

## Materials and Methods

### Participants

The study used a convenience sample and the eligibility criteria of participants age 18 or over and resident in the UK since the beginning of COVID-19 restrictions in March 2020. Participants were recruited through open adverts on social media and through internal advertising in Bangor University School of Human and Behavioural Sciences. To achieve adequate power for testing a minimum of 92 participants were required, based on a two tailed priori power analysis of 25% effect size, 0.8 power, 0.05 error (calculated using GPower Software). Effect size used was based on previously reported increases in food intake and weight gain when under high levels of stress, both prior to and during COVID-19 restrictions [15,17]. The study protocol was performed in accordance with the declaration of Helsinki and was approved by Bangor University School of Human and Behavioural Sciences Ethics Committee (ethics approval number M08-17/18).

### Measurements

Data was collected using a bespoke online survey that was administered via Qualtrics (Qualtrics Software Company Provo, UT, USA). The survey was completed anonymously, with no data collected that could identify participants. The survey was designed to be easy to answer and completed in as short a time as possible, to ensure minimal drop out of participants. To enable this, where appropriate, questions were asked about categorical changes (i.e., increased, stayed the same or reduced) rather than absolute measures. As well as simplifying the questionnaire, the use of categorical measures, rather than absolute measures, enabled the reporting of overall trends while avoiding inaccuracy problems with self-report measures. After providing informed consent, eligible participants reported on the following information:

#### Demographics and Weight Change

Participants reported on the demographic measures on age, gender, main current occupation, country of the UK based, height and weight (with BMI being calculated by the researcher). Participants were asked to report how their weight had changed since the beginning of the COVID-19 restrictions within the following categories: lost more than 3kg/6 pounds, lost up to 3kg/6 pounds; weight remained stable, gained up to 3kg/6 pounds, gained between 3kg/6 pounds and 6kg/a stone, gained more than 6kg/a stone.

#### Eating Behaviours

Participants were asked to report whether their general hunger, food cravings, number of meals consumed, snacking in the daytime and snacking in the evening had decreased, increased or stayed the same in the past 3 months, compared to prior to COVID-19 restrictions. Participants also reported if they believed any of the following reasons had increased their dietary consumption: being at home, boredom, stress anxiety or low moods, feeling tired or lacking in energy, The behaviour categories were based on categories reported by other COVID-19 studies [20,47].

#### Dietary Consumption

Participants were asked to report if their consumption of any of the following categories had decreased, increased or remained the same in the past 3 months, compared to before the start of the COVID-19 restrictions: fruit and veg; high fibre foods (such as wholemeal food, wholegrains and beans), red or processed meat (including bacon, ham, sausages; white meat (including fish), dairy products, vegetarian/vegan meat or dairy alternatives, processed foods (including ready meals or takeaways), home cooked food, tinned or frozen food; high sugar foods (such as cakes, biscuits, chocolates and sweets; high fat foods (such as savoury snacks, crisps or nuts), coffee or tea, fruit juices or smoothies, fizzy drinks (including diet drinks), alcohol, water. The range of dietary categories was based on dietary categories reported in other COVID-19 studies [15-17].

#### Physical Activity

To ensure that any reported changes in weight were due to changes in dietary consumption rather than changes in physical activity, participants were asked if their level of vigorous/moderate exercise, walking and sedentary time had decreased, increased or stayed the same in the past 3-months compared to before COVID-19 restrictions.

#### Stress

As part of the survey participants completed the Perceived Stress Scale PSS-10 [48] to evaluate general levels of stress (Cronbach’s α= 0.87, for this study). In addition, participants also reported whether prior to COVID restrictions they: had occasional stresses but generally could cope well, had some level of stress anxiety or low mood but not diagnosed, had previously been diagnosed with stress anxiety or depression, or preferred not to say.

#### Impact of COVID-19 on Lifestyle

Participants were asked to report on ways in which ways COVID-19 and the associated imposed restrictions had impacted their lifestyle and food availability. Participants were asked if the restrictions had impacted them in any of the following ways: continued to work outside the home but had to use personal protective equipment (PPE), moved to working from home, experienced job uncertainly, experienced job loss, had confirmed COVID-19 infection, had to self-isolate due to possible COVID-19 infection, had to self-isolate due to underlying health issues, had additional caring responsibilities including school-aged children or other dependents, had less access to fresh or healthy food, had less money to spend on food, had less time to prepare healthy food.

### Data Analysis

Descriptive statistics (mean ±SD) were used to characterise survey respondents and summarise their changes in weight gain, eating behaviours, dietary consumption and physical activity. Eating behaviours and dietary consumption were re-categorised in to an “increased” or “no increase” categories (which included the “stayed the same” and “decreased” categories). This was as this study was specifically interested in the likelihood of increased food cravings and comfort foods being linked to stress and weight gain.

Although the PSS-10 questionnaire gives a continuous score outcome, to enable us to carry out logistic analysis with the categorical variables reported for changes in weight, dietary consumption and exercise, participants were recategorized into a higher or lower stress group, with categorization based on the mean PSS-10 score of trial participants (19.3±6.9). Participants with a PSS score of 19.3 or lower were allocated to the lower stress category and participants with a PSS score of 19.4 or higher were allocated to the higher stress category.

Ordinal regression analyses were used to identify factors linked to a greater likelihood of reporting weight gain. Pearson’s chi-square analyses were used to investigate a significant difference in likelihood in response between the higher and lower stress groups to: weight gain, increases in eating behaviours, increases in dietary consumption, and changes in physical activity. Binary regression analysis was used to identify factors linked to a greater likelihood of belonging to the higher stress group. Odds ratios were also calculated for all chi-square analyses, to allow direct comparison of results with the binary and ordinal regression analyses. All statistical analyses were carried out using the Statistical Package for Social Sciences (IBM SPSS Statistics for Windows V.27.0. Armonk, USA). A 95% confidence interval (95% CI) was selected, and a significance level of *p* < 0.05 was applied to all statistical analyses.

## Results

### Descriptive Statistics Including Impact of COVID-19 restrictions

In total, responses were collected from 220 participants between 6th January until 2nd February 2021. After removing duplicates and incomplete questionnaires, 179 participants’ data sets remained. There was no significant demographic difference between survey completers and non-completers.

Most survey respondents were female (75%) and aged between 18-55 (85%). Two thirds of respondents were based in Wales (66%) and a third (34%) in England. The mean BMI of respondents was 26.10(±5.46) kg/m^2^.

Many respondents reported that their main current occupation involved working from home, with a higher proportion of females being based at home (F=40%:M=27%) and more male respondents continuing to work outside the home (F=15%:M=27%). As the questionnaire was advertised within the university, unsurprisingly students made up 25% of respondents, although 8% of responders were in the retired category. Notably, only females reported that caring for children or dependents was their main occupation (5%). The overall level of participants unemployed or furloughed was low (3% and 1%, respectively).

When asked about the impact of COVID-19 restrictions on their lifestyle, a comparable number of women and men reported moving to working from home (F=50%: M=46%). A higher proportion of women reported worked outside the home using PPE (F=35%:M=25%), having to isolate due to potential COVID-19 infection (F=25%:M=16%) or having additional caring responsibilities for children or other dependents (F=39%:M=9%). Males reported higher levels of job uncertainty (F=12%:M=23%), job loss (F=5%:M=7%), having had COVID-19 (F=4%:M=7%) and self-isolating due to underlying health issues (F=4%:M=11%). The difference in the level of impact of COVID-19 restrictions by gender was notable. When comparing the total number of changes that participants were affected by, 53% of females were affected by 2 or more lifestyle changes compared to 39% of males. The impact of COVID-19 restrictions on food availability was also far more likely to impact females. Females were around twice as likely to report less money to spend on food (F=16%:M=9%), less access to healthy food (F=12%:M=7%) or less time to cook healthy food (F=11%:M=5%). Full demographic statistics for survey participants are available in S1 Table.

### Stress and Weight Gain

Participants were asked to report their weight change in categories rather than absolute weight changes to avoided inaccuracy problems with self-report measures. There was a notable difference between the genders for reported weight gain, with females almost twice as likely as males to report an overall weight gain since the start of the COVID-19 restrictions (F=50%: M=27%). and twice as likely to report the highest level of weight gain of more than 6kg (F=10%: M=5%). see Table 1 for all reported weight changes.

**Table 1.** Changes in weight reported since the start of COVID-19 Restrictions.

| Weight change | Female<br>n= (%) | Male<br>n= (%) | Total<br>n= (%) |
| --- | --- | --- | --- |
| Gained more than 6kg | 13 (9.6%) | 2 (4.5%) | 15 (8.4%) |
| Gained 3-6kg | 17 (12.6%) | 3 (6.8%) | 20 (11.2%) |
| Gained up to 3kg | 37 (27.4%) | 7 (15.9%) | 44 (24.6%) |
| Weight remained the same | 40 (29.6%) | 24 (54.5%) | 64 (35.8%) |
| Lost up to 3kg | 12 (8.9%) | 1 (2.3%) | 13 (7.3%) |
| Lost more than 3kg | 16 (11.9%) | 7 (15.9%) | 23 (12.8%) |

Ordinal regression analysis showed that the likelihood of the higher stress group reporting a greater level of weight gain was statistically significant, with a medium effect size, χ^2^(5, n=179) =14.639, *p*=0.012, *phi*=0.286). This confirmed the study’s main hypothesis that participants reporting a higher level of stress during the pandemic were statistically more likely to report a higher level of weight gain. Although the aim this study was not specifically focus on gender differences, due to the large differences in weight changes reported by males and females a Chi-square tests for independence was carried out, which showed that the association between gender and weight gain was statistically significant, with small to medium effect, χ^2^(2, n=179) = 11.986, *p*=0.035, *phi*=0.259).

### Stress, Eating Behaviours and Weight Gain

In the survey, participants were asked to report on changes in eating behaviours that are often linked to weight gain, including hunger, number of meals, cravings, and snacking. Overall increases were reported for all eating behaviour categories, apart from number of meals (see Table 2). Although most respondents (53%) reported that their levels of general hunger had remained at pre-COVID-19 levels, over half of respondents reported that their food cravings, snacking in the day and snacking in the evening had increased (52%, 53% and 53%, respectively). Half of respondents also reported that the reason they had eaten more was due to being at home or boredom (52% and 48%, respectively), with 41% reporting eating more due to stress, anxiety or low mood (see Table 3).

**Table 2.** Changes in Reported Levels of Dietary Behaviours.

| Eating habits | Decreased<br>n= (%) | Stayed the same<br>n= (%) | Increased<br>n= (%) |
| --- | --- | --- | --- |
| **Eating Behaviours** |  |  |  |
| **General hunger** | 27 (15.2%) | 94 (52.8%) | 57 (32.0%) |
| **Food cravings** | 18 (10.2%) | 66 (37.5%) | 92 (52.3%) |
| **Number of meals** | 24 (13.5%) | 131 (73.6%) | 23 (12.9%) |
| **Snacking in the day** | 31 (17.4%) | 52 (29.2%) | 95 (53.4%) |
| **Snacking in the evening** | 21 (12.1%) | 61 (35.1%) | 92 (52.9%) |

**Table 3:**
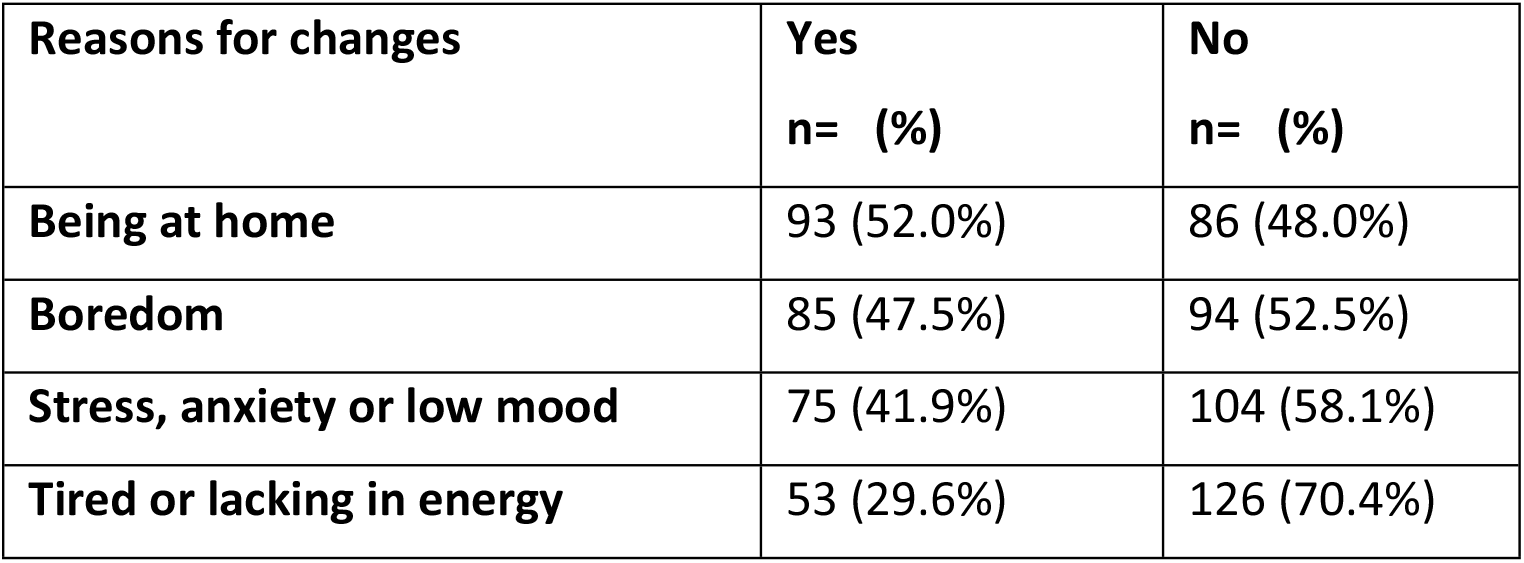
Reasons Given for Increased Food Consumption.

An ordinal logistic analysis was performed to assess whether increases in the 5 eating behaviours and 4 reasons given for increased eating increased the likelihood of respondents reporting weight gain. The full model was statistically significant, χ^2^(9, n=179) =70.877, *p*<0.001), with increased food cravings (p=0.002, OR=3.225), increased number of meals (p=0.012, OR=2.978), increased snacking in the day (p=0.012, OR=2.568) and eating due to stress, anxiety or low mood (*p*=0.026, *OR*=2.192) all significant factors linked to weight gain. Although being at home or boredom were given by the participants for the primary reasons for increased food consumption, there was no statistical significance of these factors and actual weight gain. Table 4 shows all coefficients in the ordinal logistic model.

**Table 4:**
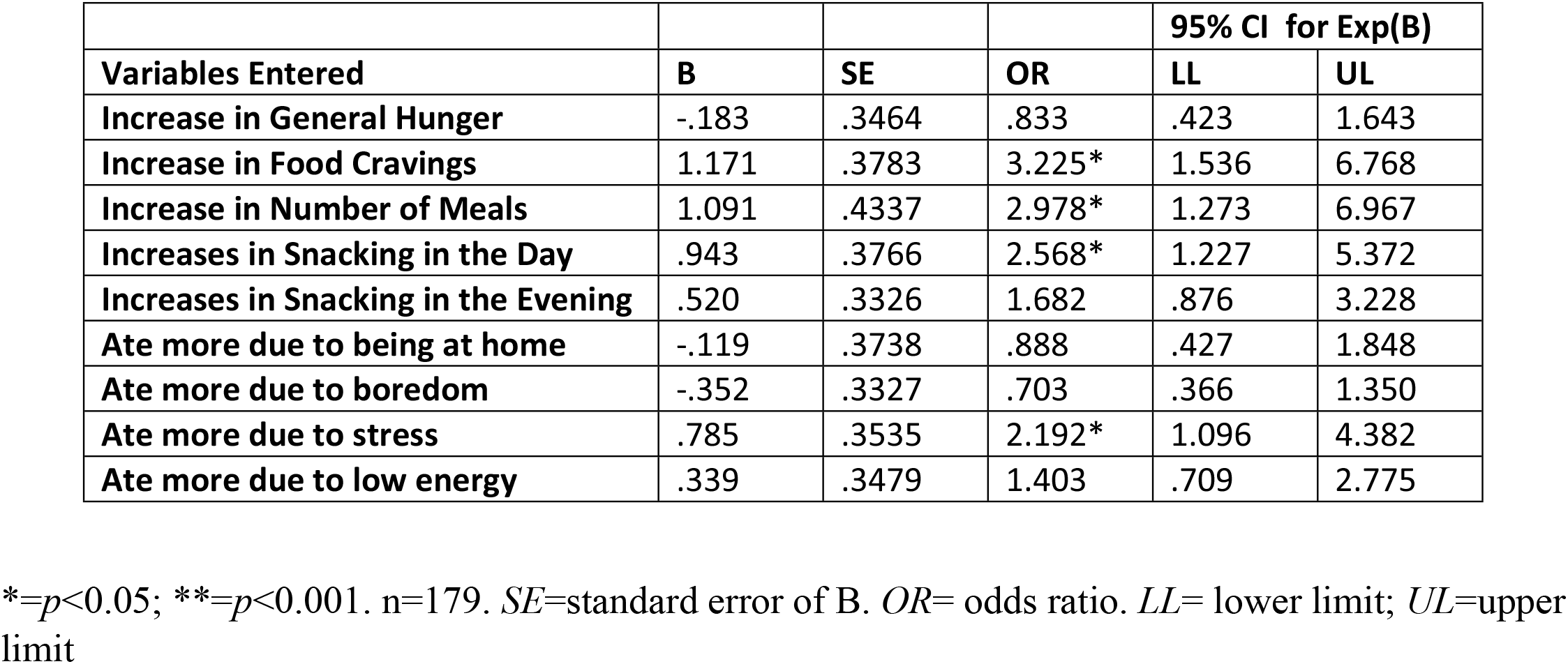
Coefficients of the predictive model for Weight Gain.

To determine whether these behaviours were more likely in the higher stress group a chi-square test was carried out. This showed that participants who were in the higher stress category were twice as likely to report increased food cravings *χ*^*2*^ (1, n=179) =6.402, *p*=0.011, *phi*=0.202;) *OR*=2.3 and snacking in the day *χ*^*2*^ (1, n=179)= 4.192, *p*=0.041, *phi*=0.165), *OR*=1.9. Participants in the higher stress category were also more likely to report increased eating due to stress, anxiety or low mood *χ*^*2*^ (1, n=179) =25.684, *p*<0.001, *phi*=0.390), which is as expected, as both measures are reporting on levels of stress. In comparison, there was no statistical significance for higher stress and increased number of meals. In summary, increased food cravings and snacking in the day were the only behaviours correlated with both higher stress and weight gain.

### Stress, Dietary Consumption and Weight Gain

Like changes in eating behaviours, there was a polarisation of changes dietary consumption. Over half of respondents maintained their pre-COVID-19 consumptions levels of fruit and veg (55%), high fibre (70%), red/processed meats (52%), white meat/fish (60%), dairy products (71%), vegetarian/vegan alternatives (62%) and tinned/frozen foods (68%). A notable change was seen with home cooked food category, where 50% increased their consumption. The other categories which showed the greatest change in consumption were those that can be categorised as comfort foods. There was comparable split between decreased, increased or remained the same for processed foods (36%, 29% and 30% respectively). 40% of participants reported an increase in consumption of high fat/salt foods, and over half (57%) of participants reported an increase in high sugar foods. With the drink categories, the majority respondents reported maintaining their pre-COVID-19 consumption level of fruit juices/smoothies, fizzy drinks and water (59%, 54% and 58%, respectively). In comparison almost half of respondents reported increased consumption of coffee and tea, and alcohol (42% and 46%, respectively). A full summary of the dietary changes for survey participants is available are available in S2 Table.

To determine which dietary categories were linked to weight gain an ordinal logistic regression was carried out. The full model, containing all 16 food and drinks categories, was statistically significant, *χ*^*2*^(16, n=179) =75.291, *p*<0.001), although only 4 of the 16 categories made a significant contribution. Increased consumption of high sugar and processed foods were linked to a 2.5-3.2-times likelihood of an increase in weight gain (*p*=0.005, *OR*=3.2 and *p*=0.032, *OR*=2.5, respectively). However, the inverse effect was seen in the categories of high fibre foods (*p*=0.034, *OR*=0.4) and water (*p*=0.033, *OR*=0.5), where increased consumption was linked to a greater likelihood of weight loss. Table 5 shows all coefficients in the ordinal logistic model.

**Table 5.** Food and Drink Coefficients of the Predictive Model for Weight Gain.

| Category | B | Std. Error | OR | Lower | Upper |
| --- | --- | --- | --- | --- | --- |
| Increase in Fruit and Veg | .445 | .3786 | 1.561 | .743 | 3.279 |
| Increase in Fibre | -1.048 | .4941 | .351* | .133 | .923 |
| Increase in Red/processed meat | -.318 | .3817 | .727 | .344 | 1.537 |
| Increase in white meat, fish and eggs | -.637 | .3901 | .529 | .246 | 1.136 |
| Increase in dairy | .850 | .4590 | 2.339 | .951 | 5.751 |
| Increase in vegetarian and vegan | -.639 | .3829 | .528 | .249 | 1.118 |
| Increase in processed/take aways/ready meals | .917 | .4274 | 2.501* | 1.082 | 5.780 |
| Increase in Home cooked | -.631 | .3431 | .532 | .272 | 1.043 |
| Increased in Tinned/Frozen | -.332 | .4978 | .717 | .270 | 1.903 |
| Increase in High Sugar foods | 1.147 | .4102 | 3.148* | 1.409 | 7.035 |
| Increase in high salt foods | .426 | .4200 | 1.531 | .672 | 3.487 |
| Increase in Coffee or Tea | .420 | .3256 | 1.521 | .804 | 2.880 |
| Increase in Juice/smoothies | .163 | .4800 | 1.177 | .460 | 3.016 |
| Increase in Fizzy drinks | .114 | .3647 | 1.120 | .548 | 2.290 |
| Increase in Alcohol | .534 | .3290 | 1.706 | .895 | 3.251 |

| Increase in Water | -.788 | .3695 | .455* | .220 | .938 |
| --- | --- | --- | --- | --- | --- |

\*= $p < 0.05$ ; \*\*= $p < 0.001$ . $n=179$ . $SE$ =standard error of B. $OR$ = odds ratio. $LL$ = lower limit; $UL$ =upper limit

A chi-square analysis was carried out to determine whether the dietary categories linked to weight change were also linked to higher levels of stress. This showed that participants in the higher stress group were twice as likely to have increased their consumption of both high sugar foods *χ*^*2*^(1, n=179) =4.001, *p*=0.045, phi=0.161), *OR*=1.9 and processed foods *χ*^*2*^ (1, n=179) = 6.310, *p*=0.012, *phi*=0.201), *OR*= 2.5. In comparison, there was no statistical significance for changes in the consumption of high fibre food or water and higher levels of stress. In summary, increases in high sugar and processed food consumption were the only dietary categories linked both to a higher level of stress and weight gain.

### Relationship between Food Cravings and Comfort Eating

To investigate further the relationship between increased food cravings and increased comfort food consumption (which includes the high sugar, high fat/salt and processed food categories), a chi-square analysis was performed. We also investigated if an increase in food cravings was linked to an increase in snacking, as comfort food is often consumed outside mealtimes [49]. Participants who reported increased food cravings were over 11 times more likely to report increased consumption of high sugar foods *χ*^*2*^(1) =48.008, *p*<0.001, *phi*=0.534; *OR*=11.2), and over 6 times more likely to report increased consumption of processed foods, *χ*^*2*^ (1) = 22.920, *p*<0.001, *phi*=0.374, *OR*=6.3), although there was no significant link with high fat/salt foods. There was also a statistically significant link between increased cravings and increased snacking in the day (but not snacking in the evening), with participants reporting increased food cravings 6 times more likely to report an increase in snacking during the day, *χ*^*2*^ (1) = 29.181, *p*<0.001, *phi*=0.419; *OR*=6.0).

### Physical Activity and weight gain

To discount the role of physical activity in the effect of stress on weight gain, participants were also asked to report in their changes in physical activity. Almost half of respondents (47%) reported a reduction in their level of moderate or strenuous activity compared to their pre-COVID-19 levels of activity. In comparison, almost half (43%) of participants reported that they had increased their level of walking compared to pre-COVID-19 level. However, the category with the greatest reported difference, was sedentary behaviour, with 78% or respondents reporting an increase in sedentary compared to their pre-COVID-19 levels. Chi-squared tests for level of activity versus weight change did show a statistical significance for a reduction in moderate/strenuous activity and weight gain, *χ*^*2*^(2, n=179) =25.354, *p*=0.005, *phi*=0.271), but not for changes in level of walking or sedentary time. Chi-squared tests of level of activity versus stress levels were non-significant *χ*^*2*^(2, n=179) =1.40, *p*=0.50, *phi*=0.088). Consequently, although a reduction of moderate/strenuous activity increased the likelihood of weight gain, it was not a mediator of the effect of stress on weight gain. A full summary of the physical activity changes for survey participants is available in S2Table. Figure 1 summarises all the significantly statistical findings reported, highlighting the corelation between higher stress, eating behaviours, dietary consumption and weight gain.

**Figure 1.**
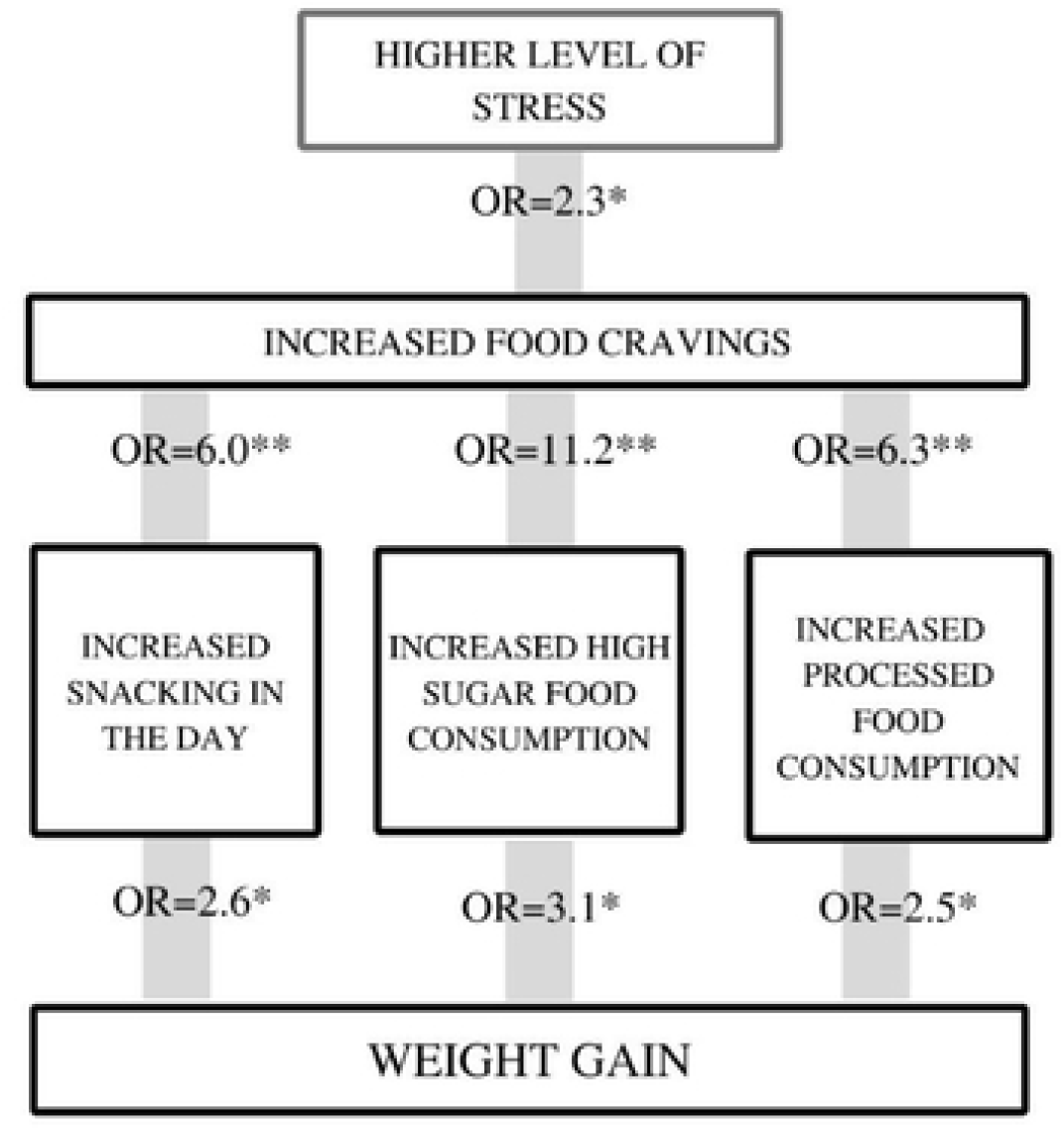
Summary of the key statistical analyses from the study. *=p<0.05; **=p<0.001. n=179. OR= odds ratio.

### Factors That Impacted Stress Levels During COVID-19

Participants were asked to report both their current levels of perceived stress, using the PSS-10 questionnaire, and their mental health prior to the start of the pandemic. With both categories a higher level of stress was reported by women. The overall PSS-10 score for trial participants was (19.3±6.9). However, the average score for women was 20.4±6.6, compared to an average score of 15.7±6.7 for men. Half of participants (54%) reported that prior to COVID-19 they had generally coped well with stress. 25% reported that they had some level of stress, anxiety or depression prior to COVID-19 restrictions, but had not received a formal diagnosis and 17% reported that they had previously been diagnosed with stress, anxiety or depression. Again, women were far more likely than men to report having been diagnosed with stress, anxiety or depression ;prior to COVID-19 (F=19%:M14%) or have had some level of stress anxiety or depression but had not been diagnosed (F=26%:M=20%).

As the pandemic caused so many enforced changes to peoples’ everyday life, one of the aims of this study was to see if any of these changes directly increased participants level of perceived stress. As reported in the descriptive statistics, moving to working from home was the highest reported change and was comparable for women and men. There were also notable differences between the genders in the numbers of changes they were affected by and some key categories such as caring from children and other dependents. A binary regression analysis was carried out which included all lifestyle changes, and gender, age and mental health prior to COVID-19. Surprisingly, the analysis did not show a significant link between any of the lifestyle changes and higher levels of perceived stress. However, gender and mental health prior to COVID-19 were both significantly linked to perceived stress during the pandemic χ2(18, n=179) =58.95, p<0.001). Participants who had a prior diagnosis of stress, anxiety or depression were almost six times more likely to be in the higher stress category (p=0.004, OR=5.7), confirming our hypothesis, with participants who had not been officially diagnosed, but reported some levels of stress, anxiety, or depression over three times more likely to be in the higher stress category (p=0.004, OR=3.6). Females were around 4 times more likely to be in the higher stress category than males (*p*=0.014, *OR*=3.6), which was independent of their levels of stress prior to COVID-19. Table 6 shows the results for all coefficients of the model.

**Table 6.**
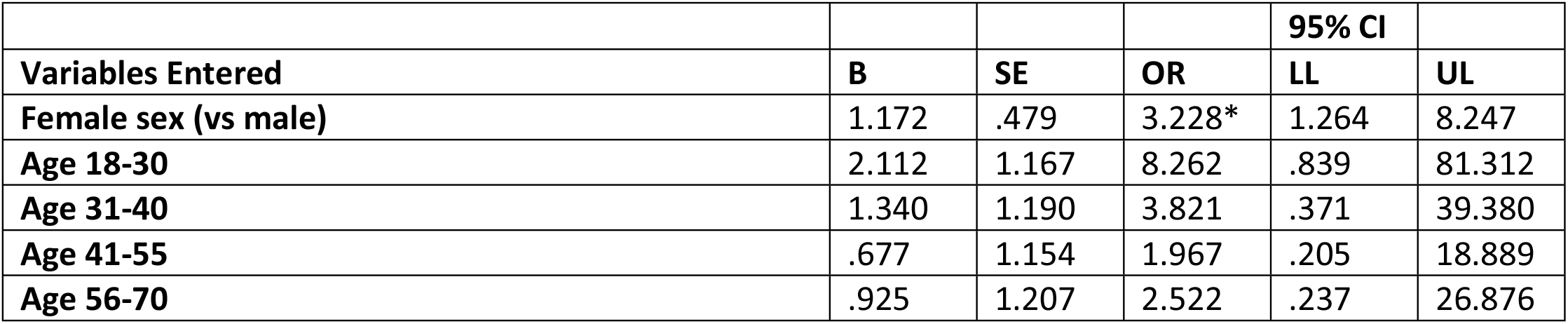
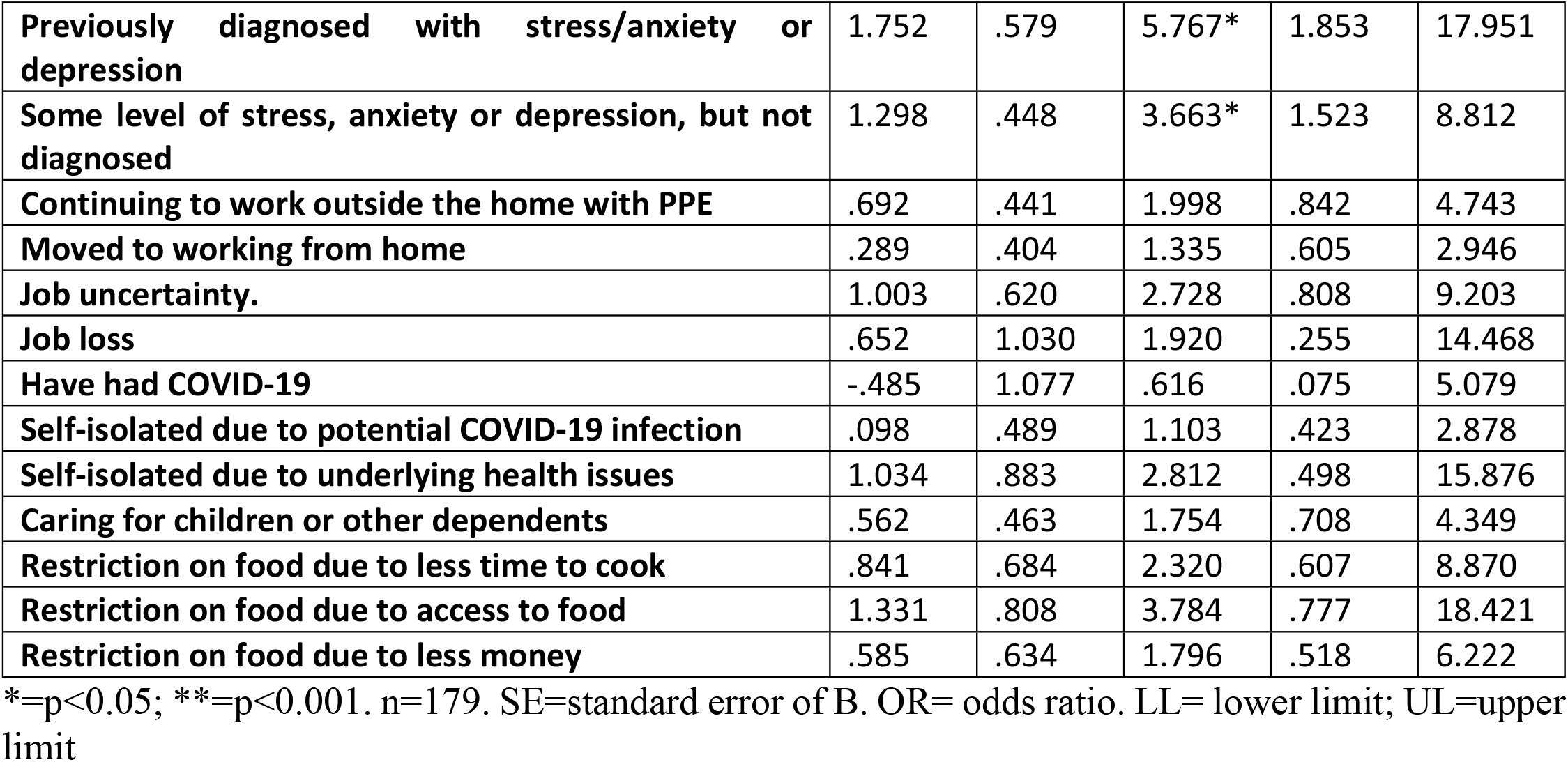
Coefficients of the predictive model for high stress (PSS score=20 or above)

## Discussion

The aim of this study was to assess whether higher levels of perceived stress during COVID-19 restrictions were linked to an increased likelihood of reporting weight gain, and whether increases in food cravings and comfort eating were mediators of this effect. We also investigated whether specific pandemic induced lifestyle changes were linked to a higher level of perceived stress and whether levels of mental health prior to COVID-19 were linked to a greater likelihood of perceived stress and weight gain.

Whilst this study was not specifically designed to investigating differences by gender, the results from the descriptive statistics revealed notable reported differences between females and males in several areas. Significantly higher stress levels were reported by females compared to males both during the pandemic and in the reporting of mental health levels prior to the pandemic. As we did not have access to PSS measurements pre-pandemic, this study is unable to say by how much perceived stress increased during the pandemic compared to pre-COVID-19 levels.

This study also reported that COVID-19 restrictions had a disproportionate impact on the everyday lives of female respondents. The higher level of working outside the home with PPE for females indicates that they were more likely to work in areas in close proximity to other people, with a likely higher risk of infection. Despite the level of COVID-19 infection being higher for men, women reported higher levels of having to isolate due to potential infection, indicating that they were in environments with a higher risk of infection, either through work or possibly due to the higher level of caring for children or dependents that they reported. This is likely to have led to increased disruption and uncertainty both for work and their personal lives. Women were also more likely to be affected by a greater number of lifestyle factors, including food availability. Although these levels of changes would be expected to cause increases in perceived stress, none of the individual lifestyle changes showed a statistically significant link with higher levels of stress. It should also be considered that specific combinations of factors or the overall number of factors may have an been required to have an impact on stress. However, this study was not designed to capture that level of complexity of analysis. It is accepted that it by using categorical rather than continuous measures for perceived stress scores we may have limited the ability to track smaller changes in lifestyle factors on stress levels. Studies highlighted in the introduction reported that stress levels during COVID-19 were reported as higher in women than in men [10-12]. As in our study, categorisation of stress was based on average PSS-10 and many females were already in the higher stress category it may be that that any additional increases in stress caused by lifestyle changes could not be detected. In former work specific indicators identified as independent factors of higher levels of stress were being female, younger and having a history of anxiety or depression [10-12]. However, as the aim of this study was to investigate the relationship between higher stress and weight gain for a wide range of categories which the using categorisation of stress levels enabled us to do.

The other area where there was a gender imbalance was in reported weight change. Overall weight gain was more likely to be reported in females than males (F=50%: M=27%) and twice as likely to be reported in females for the highest weight gain category (F=10%: M=5%). As the average BMI values of females and males at the time of the survey were comparable, it seems unlikely that this was due to long-term trends i.e., that the BMI of female respondents was already significantly higher than male respondent’s pre-pandemic. It is however acknowledged that as the results are based on self-reporting, the difference in reporting between the genders may be due to a greater awareness of weight change in females and difference in body shape satisfaction between females and males [50].

Due the number of categories studied for eating behaviours and dietary consumption, and as the study was not specifically focused on gender differences, the remaining analysis was not split by gender. Similar to previous studies our study results show a polarisation in many of the eating behaviour and dietary consumption categories, with some strong overall trends [13-18]. The main changes in eating behaviours in this study were due to increases in food cravings and snacking during the day and evening, with over half of respondents reporting increases in these categories (52%, 53% and 52%, respectively). This is in line with levels previously reported of 42-46% increase in cravings and 53-56% increase in snacking during the pandemic [20,23,47]. This study also reported that both food cravings and snacking were linked to an increased likelihood of weight gain, with increased cravings being the driver of increased snacking.

For dietary consumption, similar to previous pandemic studies [15,16] a polarisation in consumption was seen for many of the food and drinks categories. One of the main changes linked to weight gain was the change in consumption of processed foods (including take aways and ready meals) and high fat/salt foods, with increases reported of 29% and 40%, respectively. However, the greatest increase was reported was for high sugar foods, where 57% of respondents reporting an increase in consumption.

The combination of changes in behaviours and consumption are particularly concerning as this study showed that participants who reported an increase snacking or consumption of high sugar and processed foods were 2.5-3.1 times more likely to report weight gain. This appears to be driven by underlying food cravings, as participants who reported increased food cravings were 6 times more likely to increase their snacking and processed food consumption and over 11 times more likely to increase their consumption of high sugar foods. In addition, increases in food cravings were twice as likely to be reported in participants with higher levels of stress. The study highlights that food cravings and high sugar foods appear to be the main mediators of the effect of stress on weight gain. Although many previous studies have linked chronic stress to weight gain [31-34], the pandemic has given us the opportunity to observe how rapidly these behavioural pathways can be established. At the point of this study, 10 months after the initial most sever lockdowns imposed, it is apparent from our results that there was still a higher level of stress in some participants which was strongly linked to increased weight gain.

Limitations to this study are acknowledged. Although self-report surveys are useful to investigate overall trends, we acknowledge that they may not be a true reflection of actual behaviours and are often prone to bias, as they are based on responders’ perceptions of their behaviour [51]. To minimise this, questions on impact of COVID-19 restrictions and changes in eating behaviours and dietary consumption were based on previous surveys in other populations which enabled the comparison of results across similar populations. In addition, a range of general questions for eating behaviours and dietary consumption were included (rather than just asking about behaviours of interest) to limit the focus on what might perceived as negative behaviour. The study only asked about levels of change, rather than absolute measurements, so all measurements are based on comparable rather than quantitative levels of change. Perhaps the most important limitation of this work that it is a correlational study, so although we are proposing that higher stress leads to weight gain, it needs to be considered that weight gain could also be the driver of stress. We acknowledge that there are complex interactions between stress and weight gain, with different responses shown in females and males [52]. However, we proposed that by focusing on the role of food cravings this provides a stronger indication that the main direction of influence is stress, which drives food cravings, which in turn causes weight gain.

## Conclusions

In summary, the results from this study corroborate previous research, that higher levels of stress are significantly linked to increased weight gain. The study also highlights significant differences in perceived stress and weight gain during the pandemic by gender and the role in mental health prior to the pandemic in weight gain. Further work is necessary to understand how stress reduction strategies could be incorporated into weight loss/maintenance approaches, including a greater awareness of individuals’ stress levels and history of mental health when developing weight loss and maintenance strategies. A more gendered approach to weight loss may also be required, either to take into consideration differences in perceived stress levels between females and males or to determine if the role of stress in weight gain is a gendered factor.

The study also reiterates the key role of food cravings in the link between stress and weight gain. Food cravings were linked to a 3 times increased likelihood of reporting weight gain, and to 6-11 times increased likelihood of snacking and consumption of high sugar and-processed foods. This strongly suggests that re-focusing weight loss methods on the ability to control cravings, possibly by using stress reduction techniques, will be a more effective approach to than the current focus on reduction of the downstream behaviours of increased snacking, and consumption of processed and high sugar foods.

## Data Availability

The data will be held in a public repository. URL to be made available after acceptance

## Acknowledgements

The authors thank staff of the School of Human and Behavioural Sciences, College of Human Sciences, Bangor University, UK for their support and participants for taking part in the study.

## Supporting Information

S1 Table

**Descriptive statistics: demographics, reported changes due to COVID-19 and levels of stress**

S2 Table

**Reported Changes in Dietary Behaviours, Dietary Consumption and Physical Activity**

